# Frequency of *FLCN* Loss of Function Variants and Birt-Hogg-Dubé-Associated Phenotypes in a Healthcare System Population

**DOI:** 10.1101/2022.01.31.22269926

**Authors:** Juliann M. Savatt, Hermela Shimelis, Andres Moreno-De-Luca, Natasha T. Strande, Matthew T. Oetjens, David H. Ledbetter, Christa Lese Martin, Scott M. Myers, Brenda M. Finucane

## Abstract

**Purpose:** Current penetrance estimates of Birt-Hogg-Dubé (BHD)-associated cutaneous, pulmonary, and renal manifestations are based on clinically ascertained families. In a healthcare system population, we used a genetics-first approach to estimate the prevalence of pathogenic/likely pathogenic (P/LP) variants in the *FLCN* gene, which cause BHD, and the penetrance of BHD-related phenotypes.

**Methods:** Exomes from 135,990 patient-participants in Geisinger’s MyCode® cohort were assessed for P/LP *FLCN* variants. BHD-related phenotypes were evaluated from electronic health records. Association between *FLCN* P/LP variants and BHD-related phenotypes were assessed by Firth’s logistic regression.

**Results:** P/LP *FLCN* variants were identified in 35 (1 in 3,234) individuals, 68.6% of whom had BHD-related phenotype(s) including: cystic lung disease (65.7%); pneumothoraces (17.1%); cutaneous manifestations (8.6%); and renal cancer (2.9%). Only four (11.4%) were clinically diagnosed with BHD.

**Conclusion:** In this healthcare population, the frequency of P/LP *FLCN* variants is 60 times higher than the previously reported prevalence. Although most variant-positive individuals had BHD-related phenotypes in the EHR, a minority were clinically diagnosed with BHD, likely because cutaneous manifestations, pneumothoraces, and renal cancer were observed at lower frequencies than clinical cohorts. Improved clinical recognition of cystic lung disease and education concerning its association with *FLCN* variants could prompt evaluation for BHD.

## Introduction

First described in the 1970’s, Birt-Hogg-Dubé syndrome (BHD; OMIM 135150) is an autosomal dominant condition associated with benign cutaneous manifestations, pulmonary cysts/blebs/bullae (hereafter referred to as “cystic lung disease”), spontaneous pneumothoraces, and renal neoplasms.^1-2^ BHD is caused by germline variants in the folliculin (*FLCN*) gene, a tumor suppressor on chromosome 17p11.2 (OMIM 607273), that encodes the folliculin protein that interacts with numerous signaling pathways.^3-6^ Variants resulting in premature termination of the folliculin protein are the most common variant type identified in individuals with BHD.^2^

BHD has an estimated prevalence of 1 in 200,000^7^, and, as of 2017, over 600 families with BHD have been described in the literature.^8^ The most common phenotypic features in individuals with BHD are benign cutaneous manifestations, including fibrofolliculomas and trichodiscomas. Such findings have been identified in 58-90% of clinically ascertained patients and typically appear after age 25.^9^ Macroscopically, trichodiscomas and fibrofolliculomas are indistinguishable from one another and are believed to be on the same morphological spectrum along with perifollicular fibromas.^10^ Acrochordons have also been described in patients with BHD, but these lesions are also common in the general population and their relationship to BHD is unclear.^9^

Patients with BHD also have increased risk of developing cystic lung disease and spontaneous pneumothoraces. Pulmonary cysts have been identified in 67-90% of patients with BHD^2, 29,11^ Cysts vary in number and are typically bilateral and located in the lower basal zones of the lungs.^9^ The literature suggests that 24-38% of patients with BHD experience a spontaneous pneumothorax with a median age of 38 and earliest age of seven years.^2,11^

In addition to the lung and skin findings, patients with P/LP *FLCN* variants are at an increased risk for renal neoplasia. Bilateral, multifocal renal tumors are typical and are histologically diverse with most being hybrid oncocytic tumors, chromophobe renal cell carcinoma, and clear cell carcinoma.^2^ It has been estimated that 12 to 34% of individuals with BHD develop renal cancers^2, 9,11^ with a mean age of diagnosis of 50.7 years and earliest diagnosis at age 20.^9^ Additionally, individuals with BHD are at risk of developing oncocytosis, microscopic foci of oncocytic cells in the renal parenchyma, that is a precursor lesion to malignant tumors.^2^ Other phenotypes have been reported in patients with BHD including lipomas, parathyroid adenomas, thyroid nodules, thyroid cancer, and parotid oncocytomas, but additional data are needed to determine if these are associated with BHD.^2^

To date, penetrance estimates of BHD-related features in individuals with P/LP *FLCN* variants have been based on families clinically ascertained due to a personal and/or family history. Population-based testing for pathogenic variants in other disease-associated genes has suggested that clinically ascertained penetrance estimates might be higher than penetrance in variant-positive individuals from broader cohorts.^12,13^ Here, we sought to explore the prevalence of P/LP *FLCN* variants in a health care system-based population and utilize electronic health record (EHR) data to investigate the frequency of BHD-related phenotypes and prior clinical identification of the *FLCN* variant in individuals with such variants.

## Materials and Methods

### MyCode Participants

The Geisinger MyCode^®^ Community Health Initiative (MyCode) serves as a biobank of blood and other samples from over 290,000 patient-participants who consent to health-related research.^14-16^ Participants are recruited throughout Geisinger regardless of disease or phenotype.^14-17^ The population for this study includes a subset of MyCode patient-participants, referred to as the DiscovEHR cohort^14^, with available exome sequencing and linked EHR data. Most patient-participants also have available SNP genotyping data. MyCode and the research outlined below are governed by the Geisinger Institutional Review Board; all participants in this study provided written informed consent.

### Exome Sequencing and Intragenic CNV Calling

Exome sequencing for 135,990 adult MyCode patient-participants was performed in collaboration with the Regeneron Genetics Center, as described previously.^14-15^ Briefly, exome sequence data were aligned to human reference GRCh38 with BWA-mem^18,^ and the resultant BAM files processed using the Picard MarkDuplicates tool (http://broadinstitute.github.io/picard) to flag duplicate reads. After exome data were aligned, single nucleotide variants (SNVs) and insertion/deletion variants were identified using weCall.^19^ Joint genotyping was performed across the entire cohort using GLnexus.^20^ CNV calling was carried out using Copy number estimation using the Lattice-Aligned Mixture ModelS (CLAMMS) algorithm^21^ and CNV calls were validated with SNP-array CNV calls using Penn CNV.^22^

### *FLCN* Variant Analysis

We reviewed exome sequencing data (n=135,990) for potential loss of function variants in the *FLCN* gene. Variants in VCF files were annotated with Ensemble Variant Effect Predictor (VEP) version 96 using RefSeq transcripts. High impact variants (nonsense, frameshift, +/-1,2 splice site) based on the *FLCN* transcript most expressed in adult tissues (transcript variant 1, GenBank: NM_144997.7, 14 exons total with 11 coding exons, 579 amino acid residues) in genome build GRCh38.^5^ Sequence variants with an allele balance greater or equal to 0.20, genotype quality greater than or equal to 80, coverage greater than or equal to 20, and GnomAD allele frequency less than 0.001 were identified. Two *FLCN* variants that nearly met our quality thresholds were included in our analysis upon confirmation of the *FLCN* variant from a clinical genetic testing report documented in the EHR (n=1) and validation of a variant by evaluation of IGV plots (n=1). Variants were then reviewed (JMS and HS) using the American College of Medical and Genomics and Association for Molecular Pathology variant interpretation guidelines and classifications were confirmed by a third reviewer (NTS), who is board certified in laboratory genetics and genomics.^23-25^

### Determination of Relatedness

To report on the number of unique families with P/LP *FLCN* variants, the frequency of BHD-related phenotypes within families, and P/LP variant prevalence in an unrelated subset of the MyCode cohort, first- and second-degree familial relationships were identified from genotype data using Pedigree Reconstruction and Identification of the Maximally Unrelated Set (PRIMUS).^26^

### Phenotype Evaluation – EHR and Imaging Review

We characterized BHD-related pulmonary, cutaneous, and renal phenotypes in variant-positive individuals via manual review of patients’ EHRs. A chart review guide was developed to collect variables of interest and independent, double reviews were completed by a genetic counselor and board-certified neurodevelopmental pediatrician (JMS, SMM) between March and November 2020 (Supplemental Materials and Methods). All discrepancies between reviewers were resolved through joint review and consensus.

Cystic lung disease and renal masses might be incidentally identified on imaging studies of adjacent body parts (e.g., Computed Tomography (CT) of the abdomen includes the lung bases and lumbar spine Magnetic Resonance Imaging (MRI) includes at least part of the kidneys). Additionally, small lung cysts, blebs, and bullae may be identified, but sometimes not mentioned, in radiology reports or elsewhere in the EHR. Because of this, dedicated review of imaging studies including all or part of the kidneys and lungs was completed by a board-certified radiologist (AMDL) for all patients with available imaging (Supplemental Materials and Methods). Plain X-rays were not included in this review.

### Phenotype Evaluation – ICD-9/10 Diagnostic Codes

While chart review provides more accurate phenotyping, it is low throughput and often is not feasible for large sample sizes. As such, clinical manifestations of BHD-related phenotypes were also evaluated in variant positive and negative patient-participants by extracting relevant International Statistical Classification of Diseases and Related Health Problems 9 and 10 codes (ICD-9/10) from the EHR in February 2021. Selected ICD-9/10 diagnostic codes were defined prior to manual chart review and were intended to capture BHD-related pulmonary, cutaneous, and renal phenotypes (Supplemental Table 1). While ICD-9/10 codes can provide an estimate of BHD-related phenotypes, some codes that correspond to BHD-features are non-specific and could be used for other, unrelated clinical findings (e.g., ICD-10 codes for trichodiscoma and fibrofolliculoma are broadly used for benign neoplasms of the skin), and others might not be applied even when such a feature is identified due to its indolent or benign nature (e.g., ICD-9/10 codes for pulmonary cyst). Since use of ICD-9/10 codes alone might not correctly reflect patient phenotypes,^27^ we evaluated the ability of ICD-9/10 codes in the EHR to accurately identify BHD-related phenotypes by comparing diagnoses captured using diagnostic codes to those captured with manual review of EHR data in variant-positive individuals.

### Statistical Analysis

Continuous variables are summarized as median and interquartile range and categorical variables are described as frequency and percentage. Fisher’s exact and Wilcoxon rank-sum tests were performed to test significant differences in demographics between variant-positive and variant-negative individuals. Associations between *FLCN* P/LP variants and BHD phenotype were assessed by comparing the frequency of BHD-related phenotypes in variant-positive individuals with the variant-negative group using relevant ICD-9/10 diagnostic codes.The statistical significance of associations between *FLCN* P/LP variants and BHD-related phenotypes was calculated using Firth logistic regression model adjusting for age, sex and the first four principal components of ancestry. To reduce the impact of population stratification on the results, the European subset of the cohort (121,876) was used for the association analysis. The European subset used in the association analysis was identified by principal component analysis and clustering with the 1000 Genomes reference population using the same protocol applied to the UK Biobank.^29^ Bonferroni correction was used to account for multiple testing(α = 0.05). All analyses were conducted using R version 4.0.1.

## Results

### Prevalence of P/LP *FLCN* Variants

Of the 135,990 patient-participants, 35 (0.03%, 1 in 3,234) had a P/LP variant in *FLCN*. Based on genetic relatedness data,^26^ we determined that these 35 individuals are from 28 families, representing 0.03% of the 90,563 first and second-degree unrelated subset of the cohort. Thirteen unique *FLCN* variants were identified, including five frameshift variants identified in 17 individuals, five nonsense variants identified in six patient-participants, two canonical splice site variants identified in 12 individuals, and a single intragenic deletion of exons 4-11 in one participant (Figure 1). One individual had two variants identified -- deletion of exons 4-11 and a frameshift variant in cis. Nine of the thirteen variants identified in 32 of the 35 participants were previously reported in the literature and/or submitted to the National Center for Biotechnology Information ClinVar^29^ database. Table 1 summarizes demographics of *FLCN* variant-positive and variant-negative individuals in the MyCode cohort. Patient-participants with *FLCN* variants were 62.9% female, 97.1% self-reported their race as White, 100% self-reported non-Hispanic ethnicity, and had a median age of 62 years. Eighty-nine percent of patient-participants were alive, and the median length of EHR data was 17.43 years at the time of manual chart review. No statistically significant demographic differences were identified between the variant-positive and variant-negative individuals.

**Table 1.**
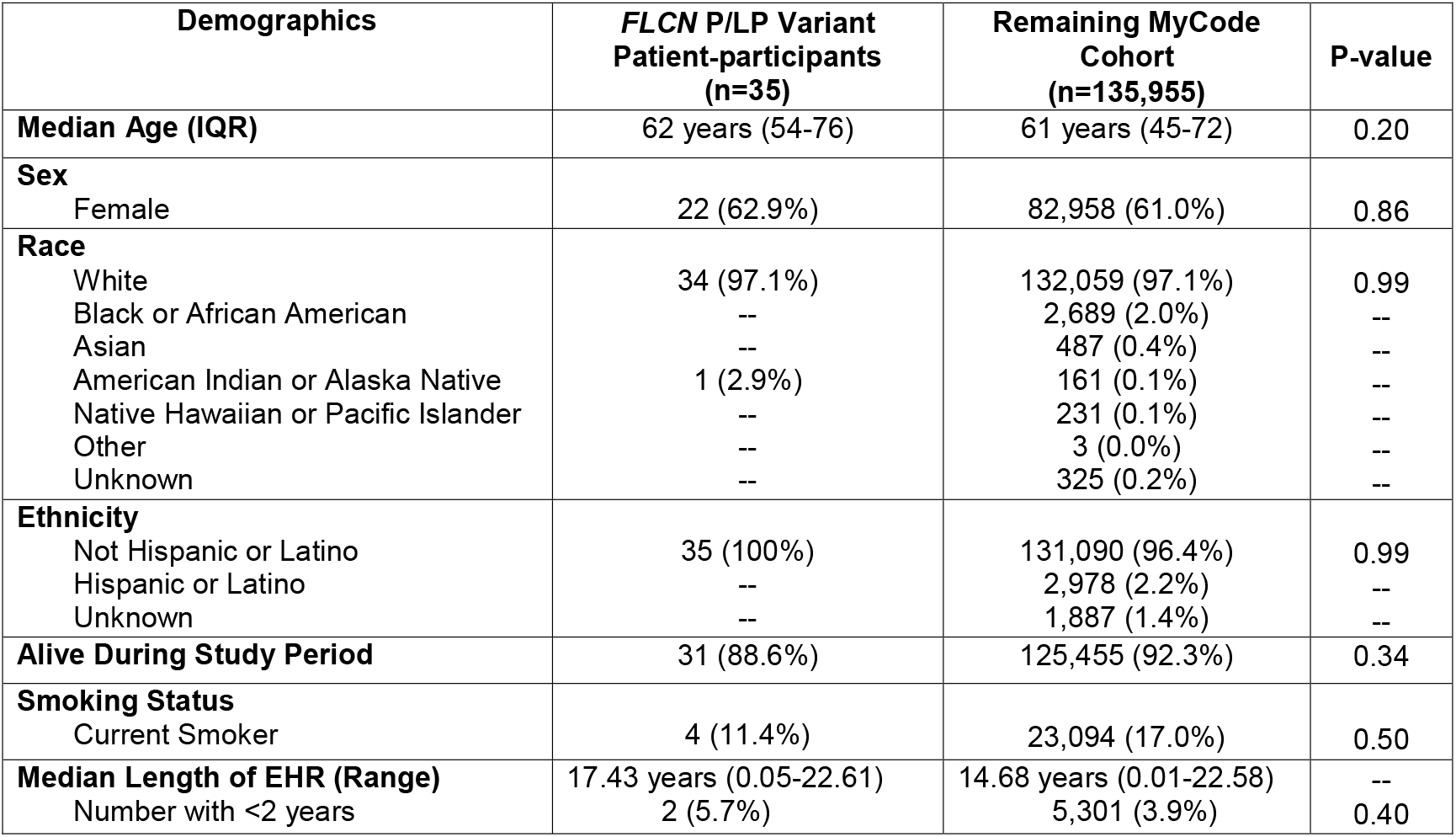
Demographic Details in *FLCN* Variant Positive Patient-Participants and the Overall MyCode Cohort.

| **Demographics** | ***FLCN* P/LP Variant**<br>**Patient-participants**<br>**(n=35)** | **Remaining MyCode**<br>**Cohort**<br>**(n=135,955)** | **P-value** |
| --- | --- | --- | --- |
| **Median Age (IQR)** | 62 years (54-76) | 61 years (45-72) | 0.20 |
| **Sex** |  |  |  |
| Female | 22 (62.9%) | 82,958 (61.0%) | 0.86 |
| **Race** |  |  |  |
| White | 34 (97.1%) | 132,059 (97.1%) | 0.99 |
| Black or African American | -- | 2,689 (2.0%) | -- |
| Asian | -- | 487 (0.4%) | -- |
| American Indian or Alaska Native | 1 (2.9%) | 161 (0.1%) | -- |
| Native Hawaiian or Pacific Islander | -- | 231 (0.1%) | -- |
| Other | -- | 3 (0.0%) | -- |
| Unknown | -- | 325 (0.2%) | -- |
| **Ethnicity** |  |  |  |
| Not Hispanic or Latino | 35 (100%) | 131,090 (96.4%) | 0.99 |
| Hispanic or Latino | -- | 2,978 (2.2%) | -- |
| Unknown | -- | 1,887 (1.4%) | -- |
| **Alive During Study Period** | 31 (88.6%) | 125,455 (92.3%) | 0.34 |
| **Smoking Status** |  |  |  |
| Current Smoker | 4 (11.4%) | 23,094 (17.0%) | 0.50 |
| **Median Length of EHR (Range)** | 17.43 years (0.05-22.61) | 14.68 years (0.01-22.58) | -- |
| Number with <2 years | 2 (5.7%) | 5,301 (3.9%) | 0.40 |

**Figure 1.**
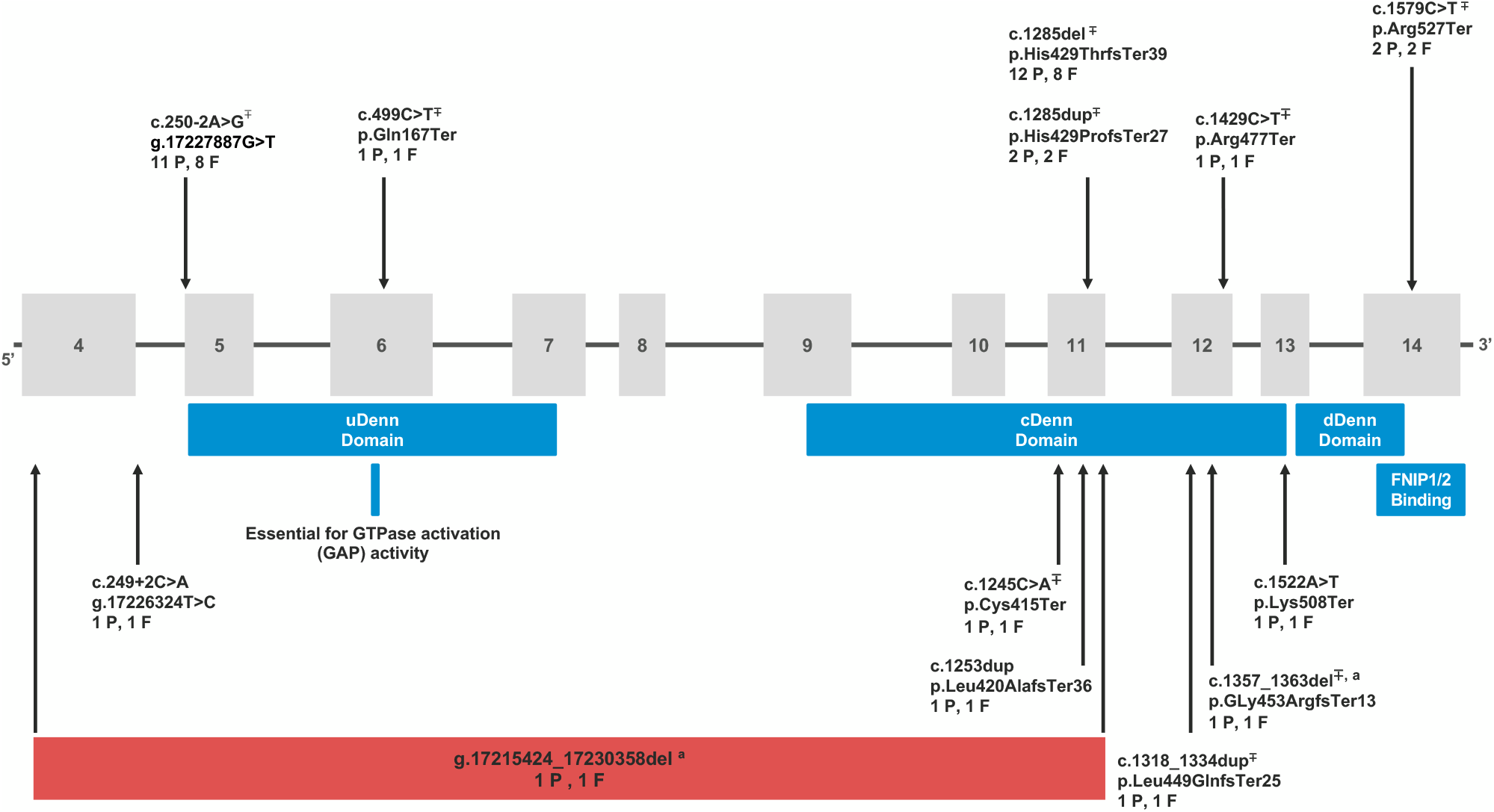
Pathogenic/Likely Pathogenic variants in *FLCN* identified in the MyCode cohort. Based on build GRCh38 (NC_000017.11) and *FLCN* transcript most expressed in adult tissues (transcript variant 1, GenBank: NM_144997.7, 14 exons total with 11 coding exons, 579 amino acid residues) P=number of patient-participants with that variant and F=number of families with that variant. ∓denotes variants that were previously reported in the literature and/or submitted to ClinVar. ^**a**^ denotes two variants identified in the same participant.

### BHD-related Phenotypes – Chart Review of Patient-participants with *FLCN* Variants

The frequency of BHD-related phenotypes (i.e., cutaneous, pulmonary, and renal manifestations) was evaluated by manual chart review of each variant-positive patient-participant. We found that 68.6% (n=24/35) of variant-positive individuals had EHR-documentation of a BHD-related phenotype (Figure 2 and Table 2).

**Table 2.**
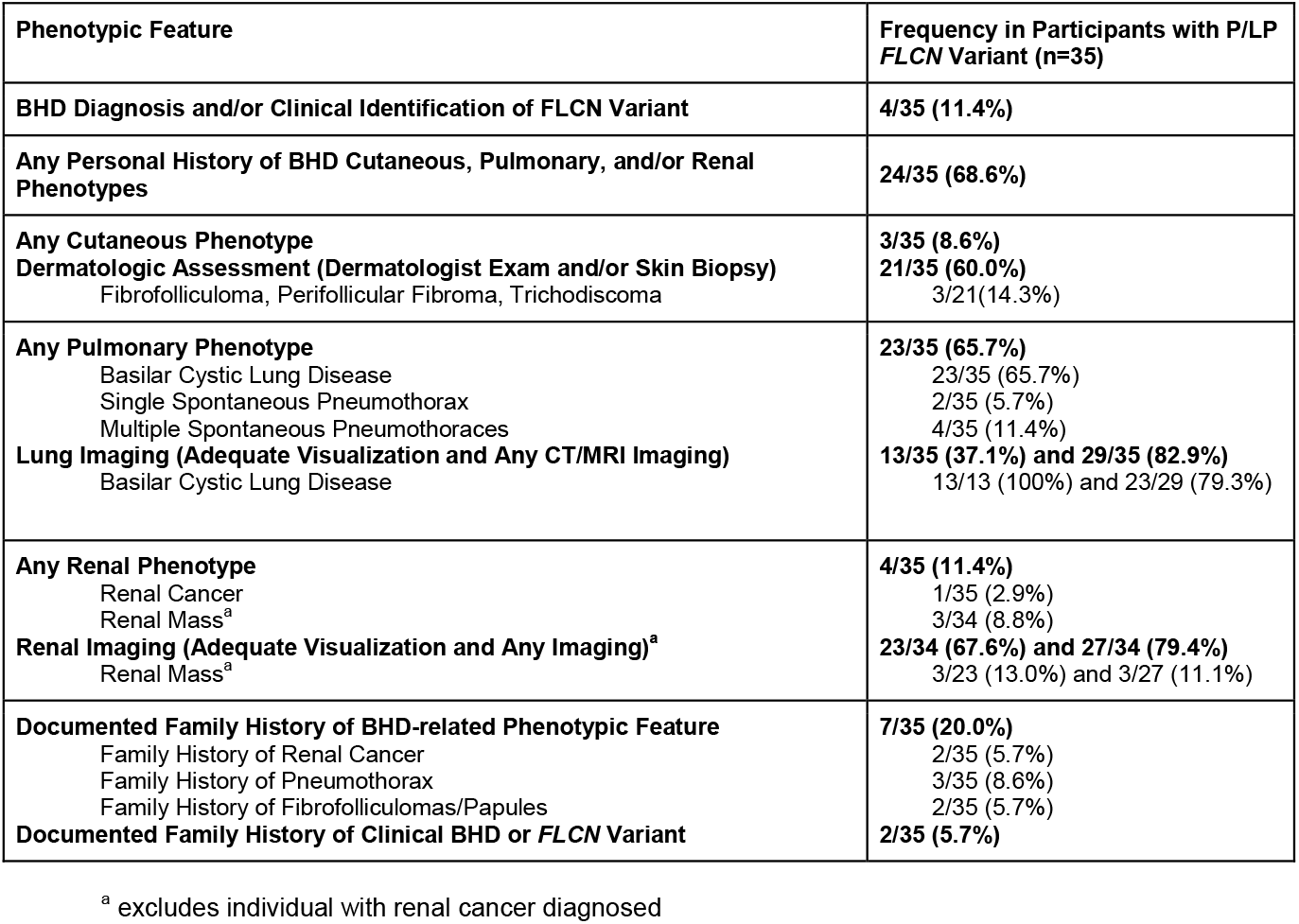
EHR Review for BHD-related Phenotypic Features in Variant Positive Patient-Participants.

**Figure 2.**
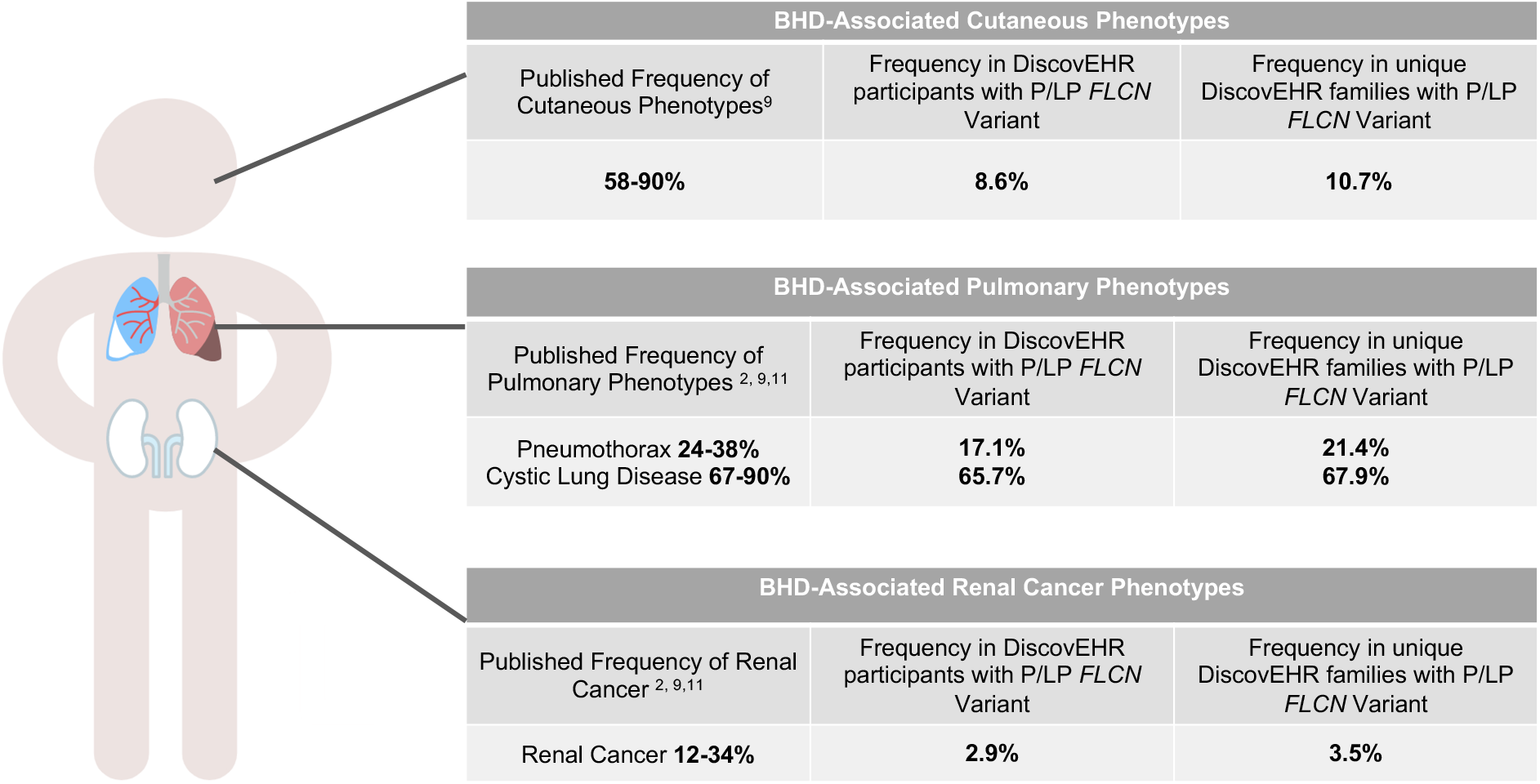
Frequency of BHD-related phenotypes in the literature compared to that in individual MyCode patient-participants with P/LP *FLCN* variants and in unique MyCode families. Exome data was used to identify first-second degree relationships and to group individuals into unique families.

BHD-related cutaneous findings including fibrofolliculomas, perifollicular fibromas, and trichodiscomas were identified in three patient-participants (8.6%). Among all variant-positive participants, 60% (n=21/35) had documentation of an evaluation with a dermatologist and/or at least one skin biopsy. When restricting to those who were examined by a dermatologist and/or had a skin biopsy 14.3% (n=3/21) had a BHD-related cutaneous finding. BHD-related cutaneous findings were identified in 10.7% (n=3/28) of unique families with *FLCN* variants.

Pulmonary phenotypes, including basilar cystic lung disease and/or pneumothorax, were identified in 65.7% (n=23/35) of patient-participants. Of the 23 individuals with pulmonary phenotypes, six had at least one spontaneous pneumothorax; two had exactly one and four had more than one. Participants experienced their first pneumothorax between the ages of 21 and 79 years (median 27.5 years). The individual that experienced their first pneumothorax at age 79 had a diagnosis of chronic obstructive pulmonary disease. All 23 individuals with a pulmonary phenotype had basilar cystic lung disease. Eighty-three percent (n=29/35) of variant-positive patient-participants had CT or MRI imaging that included at least a portion of the lungs (e.g., abdomen CT includes lung bases) and 37.1% (n=13/35) had adequate visualization of the entire lungs. Among those with any lung imaging and those with adequate imaging reviewed, 79.3% (n=23/29) and 100% (n=13/13) had basilar cystic lung disease, respectively. Among unique families with *FLCN* variants, 21.4% (n=6/28) had pneumothorax and 67.9% (n=19/28) had cystic lung disease in at least one family member (Figure 2).

Renal cancer was identified in one patient-participant (2.9%) with a chromophobe renal cancer diagnosed at age 71. Among the remaining 34 patient-participants without renal cancer, 79.4% (n=27/34) had imaging that included the kidneys and 67.6% (n=23/34) had adequate visualization of both kidneys in their entirety. Among those with any renal imaging and those with adequate renal imaging (e.g., bilateral renal ultrasound), 11.1% (n=3/27) and 13.0% (n=3/23) had renal masses identified, respectively. These renal masses had not been pathologically confirmed to be renal cancer.

In addition to core BHD-related phenotypes, the EHR review included evaluation of other phenotypes that have been reported in patients with BHD including thyroid cancer and parotid oncocytomas. Bilateral parotid oncocytosis was noted in a single participant with bilateral cystic lung disease. Thyroid cancer was not noted in any individuals.

Four individuals (11.4%) from three families had a diagnosis of BHD or documentation of a germline *FLCN* variant in their EHR. Two of those individuals had EHR documentation of multiple spontaneous pneumothoraces and bilateral blebs; one had a fibrofolliculoma, bilateral blebs, and renal mass; and one had a perifollicular fibroma.

### BHD-related Phenotypes – Diagnostic Code Comparison

Comparison of BHD-related phenotypes captured by ICD codes with manual chart review (Supplemental Table 2) revealed that ICD-9/10 codes for BHD-related cutaneous features lack specificity and, as such, estimate the rates of such findings in variant-positive individuals at a higher rate than observed via manual review. We found that 42.9% (n=15/35) versus 8.6% (n=3/35) had BHD-related cutaneous features based on the ICD code approach and chart review respectively. ICD codes underestimated the frequency of cystic lung disease [20.0% (n=7/35) based on ICD codes versus 65.7% (n=23/35) chart and radiology review], likely due to the benign nature of these findings and incidental identification on imaging. Diagnostic codes for pneumothorax were found to accurately identify this phenotype in variant-positive individuals [17.1% (n=6/35) ICD codes versus 17.1% (n=6/35) chart and radiology review]. All six individuals with pneumothoraces identified on manual chart review had an appropriate ICD-9/10 code in their EHR. ICD diagnostic codes for renal cancer appropriately identified the single individual with renal cancer. Diagnostic codes associated with a renal mass underestimated this phenotype [2.9% (n=1/34) ICD codes versus 8.8% (n=3/34) chart review].

The associations between *FLCN* P/LP variants and ICD-9/10 based BHD-related phenotypes were assessed by Firth’s logistic regression in the unrelated, European subset of the MyCode cohort (Table 3). P/LP variants in *FLCN* were associated with a higher prevalence of pulmonary phenotypes (OR=4.16; 95% CI:11.65-9.43; p=0.026), and, more specifically, spontaneous pneumothorax (OR=15.63; 95% CI: 5.96-35.69; p=1.92×10^**−**5^) in variant positive patient-participants. Association of *FLCN* P/LP variants with other pulmonary, cutaneous, and renal phenotypes were not statistically significant when correcting for multiple testing. We performed the association analysis in the full unrelated subset of the cohort including European and non-European minority populations, to increase sample size. The results of the full unrelated cohort were consistent with those of the European subset (Supplemental Table 3).

**Table 3.**
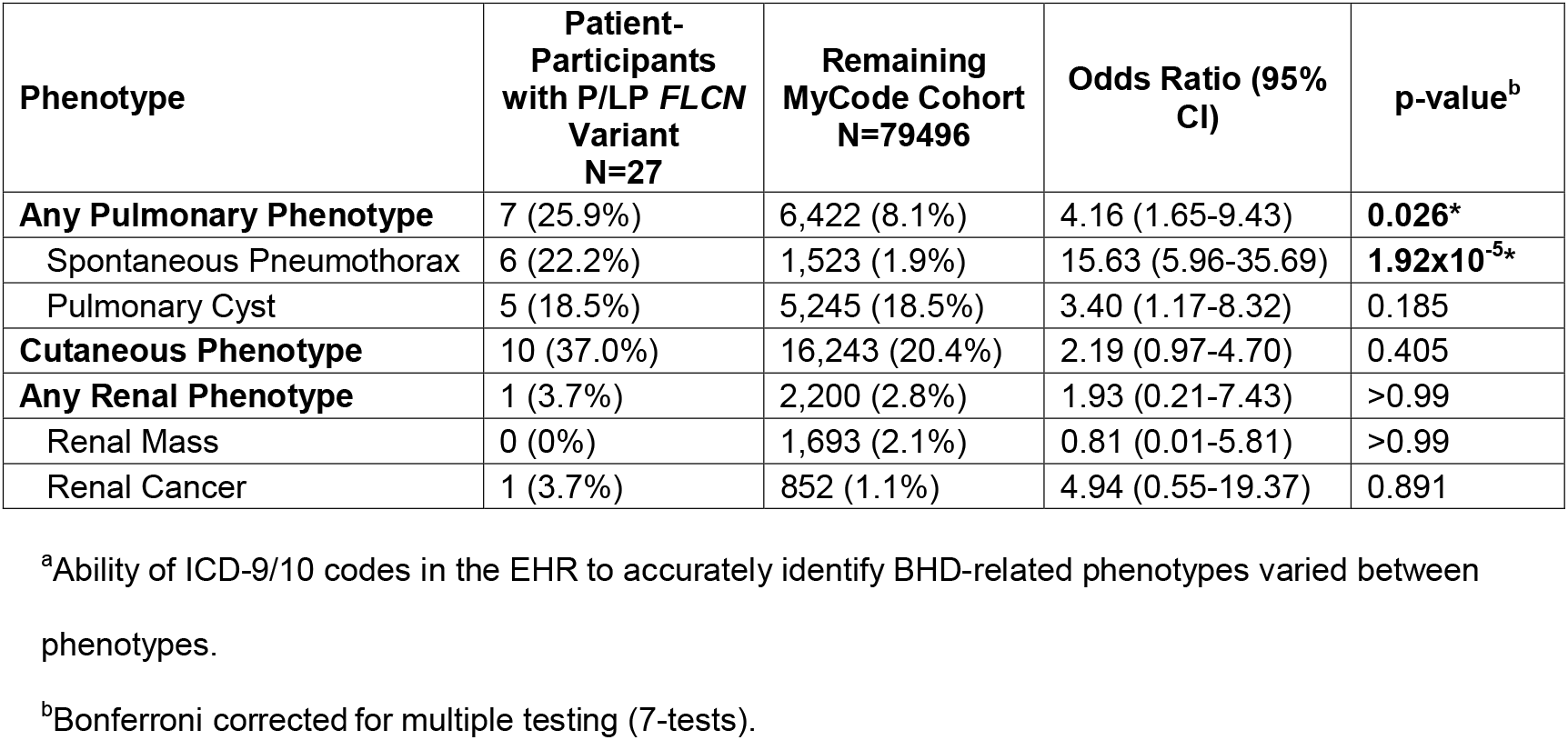
**Associations Between *FLCN* P/LP Variants and Diagnosis of BHD-related Phenotypes^a^ in the First- and Second-Degree Unrelated European Subset of the MyCode Cohort**

| Phenotype | Patient-Participants with P/LP *FLCN* Variant N=27 | Remaining MyCode Cohort N=79496 | Odds Ratio (95% CI) | p-value<sup>b</sup> |
| --- | --- | --- | --- | --- |
| **Any Pulmonary Phenotype** | 7 (25.9%) | 6,422 (8.1%) | 4.16 (1.65-9.43) | **0.026\*** |
| Spontaneous Pneumothorax | 6 (22.2%) | 1,523 (1.9%) | 15.63 (5.96-35.69) | **1.92x10<sup>-5</sup>\*** |
| Pulmonary Cyst | 5 (18.5%) | 5,245 (18.5%) | 3.40 (1.17-8.32) | 0.185 |
| **Cutaneous Phenotype** | 10 (37.0%) | 16,243 (20.4%) | 2.19 (0.97-4.70) | 0.405 |
| **Any Renal Phenotype** | 1 (3.7%) | 2,200 (2.8%) | 1.93 (0.21-7.43) | >0.99 |
| Renal Mass | 0 (0%) | 1,693 (2.1%) | 0.81 (0.01-5.81) | >0.99 |
| Renal Cancer | 1 (3.7%) | 852 (1.1%) | 4.94 (0.55-19.37) | 0.891 |

<sup>a</sup>Ability of ICD-9/10 codes in the EHR to accurately identify BHD-related phenotypes varied between phenotypes.
<sup>b</sup>Bonferroni corrected for multiple testing (7-tests).

## Discussion

### Frequency of P/LP *FLCN* Variants

BHD is considered a rare disorder with an estimated prevalence of 1 in 200,000.^7^ BHD prevalence and penetrance estimates have been based on clinically ascertained families to date. This study examined exome sequencing data from a large health care population to estimate the prevalence of P/LP *FLCN* variants and frequency of BHD-related phenotypes.*FLCN* variants were identified in 28 out of 90,563 first- and second-degree unrelated individuals (1 in 3,234) suggesting that such variants are significantly more common than previously reported. This finding is consistent with the increased frequency of variants associated with other genetic syndromes identified in broader, population cohorts via genome-first approaches.^30-32^

### Prevalence of BHD-related Phenotypes

In participants with P/LP *FLCN* variants from this healthcare cohort, BHD-related lung findings were the most common phenotype identified (65.7%, n=23/35). The frequency of basilar pulmonary cystic lung disease in those with CT and MRI lung imaging (79.3%, n=23/29) overlapped with the reported frequency in clinically ascertained individuals (67-90%)^2, 9,11^ and was even higher than the reported frequency in those with adequate visualization of the lungs (100%). The frequency of pneumothorax (17.1%, n=6/35) was lower than that reported in the published literature (24-38%). Pneumothorax-associated ICD-9/10 diagnostic codes were present in the EHR of all patient-participants with pneumothoraces identified by chart review, suggesting that these codes capture the phenotype with high sensitivity. When we assessed the association between *FLCN* P/LP variants and pneumothorax, we identified a higher prevalence of spontaneous pneumothorax diagnosis (OR=15.63 95%CI (5.96-35.69), p=1.92×10^**−**5^) in variant positive individuals. Together, these data suggest that while patient-participants with *FLCN* variants in this healthcare-based cohort may have lower rates of pneumothorax compared to the clinically ascertained population with BHD, they have higher rates than the variant-negative population. The prevalence of diagnosis codes associated with cystic lung disease was not statistically different between the variant-positive and the variant-negative patient-participants. However, this could be due to inability of the selected ICD 9/10 codes to identify all individuals with the phenotype, as a minority of variant-positive participants with this phenotype based on manual chart review had the relevant ICD codes in the EHR. Therefore, this pulmonary finding, which can be a precursor to spontaneous pneumothorax, may also occur at higher rates in individuals with *FLCN* P/LP variants. Additional research is needed to determine whether *FLCN* P/LP variants are associated with increased prevalence of basilar cystic lung disease.

Other BHD-related phenotypes, including cutaneous manifestations and renal cancer appeared to be less common in the variant-positive individuals in this cohort compared with BHD probands in the reported literature. BHD-related dermatologic findings were identified in only 8.6% (n=3/35) of *FLCN* variant-positive individuals compared to 58-90% of patients with BHD in the literature.^9^ When restricting the analysis to patient-participants who were examined by a dermatologist and/or had a skin biopsy that would have likely identified these benign phenotypes, only 14.3% (n=3/21) had BHD-related skin findings. This estimate is notably lower than the frequency of BHD-related dermatologic findings reported in the literature (14.3% vs. 58-90%).^9^ Since lack of clinical assessment does not alone account for differences in frequency, this finding is consistent with a lower penetrance of *FLCN* P/LP variants than previously reported. *FLCN* P/LP variants were not associated with an increased rate of cutaneous diagnosis, based on ICD-9/10 codes. Given the non-specific nature of cutaneous ICD-9/10 codes and their overestimation of BHD-related skin-findings in variant-positive individuals, however, additional assessment comparing the frequency of BHD-related cutaneous findings between variant-positive and variant-negative populations is needed.

Only one participant with a *FLCN* variant was identified to have renal cancer (2.9%, n=1/35). This is a lower frequency than reported in the literature (12-34%).^2, 9,11^ Although some participants are younger (e.g., five participants with *FLCN* variants are less than 40 years of age) and could go on to develop a renal cancer in their lifetime, the majority of *FLCN* variant-positive individuals in the cohort are over the age of 50.7 years (74.3%, n=26/35) which is the median age of renal cancer diagnosis reported in the literature for individuals with BHD. Due to the indolent nature of BHD-related renal cancers,^2^ some variant-positive patient-participants might have yet to come to clinical attention. If we assume the three participants with a renal mass noted on imaging have an undiagnosed renal cancer and only consider those with adequate renal imaging as being assessed for a renal cancer, an upper limit of potential renal cancer in variant-positive individuals is 18.2% (n=4/22). *FLCN* P/LP variants were not associated with increased prevalence of renal cancer in the MyCode cohort based on EHR data. Although additional studies are needed due to small numbers, this suggests the rates of renal cancer among individuals with *FLCN* variants may not be as high as previously estimated.

These data suggest that, among individuals in a health system population who volunteered to participate in genetic research but were not selected for any disease state, the majority of individuals with P/LP *FLCN* variants have a phenotype consistent with BHD. Individual BHD-related phenotypes including pneumothorax, cutaneous findings, and renal cancer may be less common among individuals with P/LP *FLCN* variants than previously reported, a finding that is consistent with other studies examining frequency of disease-associated phenotypes in population-based cohorts.^12,13^ Although we are not aware of any prior population-based studies of *FLCN* variants, the *FLCN* gene is located within the recurrent copy number variant region on 17p11.2 that is deleted in 90% of individuals with Smith-Magenis syndrome (SMS), hence these individuals are haploinsufficient for *FLCN*; however, there are few published reports of BHD-related phenotypes in individuals with SMS.^33-36^ Therefore, not all individuals with *FLCN* haploinsufficiency develop BHD, further suggesting that published penetrance of BHD-related phenotypes may be overestimated.

### Clinical Identification of *FLCN* Variants

Even though most participants with a P/LP *FLCN* variant had features consistent with BHD documented in the EHR, the majority of variant-positive patient-participants (88.6%, n=31/35) did not have a clinical diagnosis of BHD. The majority of those with BHD-related features had basilar cystic lung disease (95.8%, n=23/24 with phenotype). Although these features are included in the diagnostic criteria for BHD,^2,37^ their presence alone did not prompt a referral to genetics or clinical genetic testing in any patient-participants with *FLCN* variants.Even in the four individuals with clinically identified *FLCN* variants or BHD diagnoses in this cohort, all had cystic lung disease but were evaluated due to other phenotypes (fibrofolliculoma, perifollicular fibroma, pneumothoraces, family history of BHD). Although cystic lung disease was identified in 23 individuals in total, nine patient-participants were only identified on radiologist review of imaging data, meaning these findings were not documented in the EHR and were not commented on in the initial radiology report. Improved clinical recognition of these benign findings and education concerning their potential association with *FLCN* variants could prompt additional evaluation for BHD. Identifying patients at risk for BHD-phenotypes could inform appropriate management. For example, in patients with BHD, a first pneumothorax might be treated more aggressively with pleurectomy/pleurodesis given the risk for recurrent pneumothoraces^38-39^ and identification of individuals with *FLCN* variants could enable earlier screening for renal cancer, potentially allowing detection of cancers at earlier stages.^40^

### Limitations

This study has several limitations. Because the MyCode population is primarily of European ancestry and from a single healthcare system, it is unclear if these findings are generalizable to more diverse patient populations. Our estimates of BHD-related phenotypes rely on data within the EHR; as such, not all patient-participants have had assessments that would identify BHD-related features. Other studies have identified differential risks for BHD-related phenotypes depending on the *FLCN* variant.^38^ Given the small sample size and small number of recurrent variants, this analysis was not completed in this study and could be considered in future work

## Conclusions

P/LP *FLCN* variants were identified in 1 in 3,234 individuals in a population-based healthcare population. Of those with such variants, 65.6% were identified to have BHD-related phenotypes; however, only 11.4% had been clinically diagnosed with BHD. This limited ascertainment is likely because classic BHD-related phenotypes, such as cutaneous manifestations, pneumothorax, and renal cancer, were observed at lower frequencies. Cystic lung disease was the most common phenotype identified in our cohort and could be used to prompt additional evaluation for BHD. This finding could inform future screening strategies for BHD in large-scale healthcare patient populations.

## Data Availability

The data that support the findings of this study are available within the article and/or are available on request from the corresponding author, J.M.S.

## Acknowledgements

This work was supported by the National Institutes of Mental Health of the National Institutes of Health (NIH) under award number: R01 MH074090. The authors would like to acknowledge the MyCode patient-participants that make this work possible. We would also like to acknowledge those involved in the Geisinger-Regeneron DiscovEHR Collaboration who have enabled the generation of the genomics data used in this study and Dr. H. Les Kirchner for his thoughtful review and comments that helped shape the statistical analysis.

## Author Information

Funding Acquisition - C.L.M., D.H.L.; Conceptualization - J.M.S., H.S., B.M.F., S.M.M; Supervision - B.M.F., C.L.M., M.T.O., D.H.L; Project Administration - J.M.S.; Data Curation - J.M.S., H.S., S.M.M., A.M.D.L; Investigation, Methodology - J.M.S., H.S., S.M.M., A.M.D.L, N.T.S; Resources - H.S., M.T.O.; Formal Analysis - H.S.; Visualization, Writing - original draft - J.M.S., H.S.; Writing - review & editing- J.M.S., H.S., B.M.F., C.L.M., M.T.O., D.H.L, A.M.D.L., S.M.M, N.T.S.

## Ethics Declaration

The Geisinger MyCode^®^ Community Health Initiative (MyCode) serves as a biobank of blood and other samples from over 290,000 patient-participants who consent to health-related research (https://www.geisinger.org/precision-health/mycode). MyCode and the research outlined are approved by the Geisinger Institutional Review Board. MyCode participants or their parent/legal guardian provide written consent and HIPAA authorization. As part of participation, they agree to provide samples for broad research use and permit access to data in their electronic health record for research use. ^14^ Individual data included in this manuscript have been de-identified and presented in aggregate.

## Notes

### Competing Interest Statement

David H Ledbetter: Employee of Unified Patient Network, Inc., and scientific consultant for Natera, Inc., MyOme, Inc., and Seven Bridges Genomics, Inc.

### Author Declarations

The Geisinger MyCode Community Health Initiative (MyCode) serves as a biobank of blood and other samples from over 290,000 patient-participants who consent to health-related research (https://www.geisinger.org/precision-health/mycode). MyCode and the research outlined are approved by the Geisinger Institutional Review Board. MyCode participants or their parent/legal guardian provide written consent and HIPAA authorization. As part of participation, they agree to provide samples for broad research use and permit access to data in their electronic health record for research use (doi:10.1038/gim.2015.187). Individual data included in this manuscript have been de-identified and presented in aggregate.

