## Supplemental Materials and Methods for "Frequency of *FLCN* Loss of Function Variants and Birt-Hogg-Dubé-Associated Phenotypes in a Healthcare System Population"

**EHR Review Abstraction Manual for Participants with *FLCN* P/LP Variant**

**Demographics**

- Date of birth
- Deceased
- Date of death (if applicable)
- First Encounter
  - Capture the first encounter that is an office visit /in patient/surgery
- Last Encounter
  - Capture the first encounter that is an office visit/in patient/surgery

**Previous Genetic Testing**

- Search term → “*FLCN*” and “Birt Hogg Dubé”
- Lab → Filter Miscellaneous Order and look for genetic test
- If found → capture is clinical or genetic diagnosis, age of diagnosis, testing indication

**Dermatology**

- Search problem list and medical history (audit trail provides dates) → “fibrofolliculoma,” “trichodiscoma,” “perifollicular fibromatosis cutis”
- Look for Pathology Records (scans or labs) → “fibrofolliculoma,” “trichodiscoma,” “perifollicular fibromatosis cutis”
- Use text search function and look for → “fibrofolliculoma,” “trichodiscoma,” “perifollicular fibromatosis cutis”
- If found --> capture age of diagnosis or, if no age of diagnosis is clear capture the earliest age was documented. In addition to age of diagnosis, capture number identified or multiple if not quantified.
- If skin-related ICD-9/10 code was pulled automatically from the chart, look at why that code was identified.

**Lungs**

- Search Problem List for → “spontaneous pneumothorax,” “pneumothorax”
- Use text search function and look for → “Pneumothorax,” “collapsed lung,” “pneumatocele,” “lung cyst,” “pulmonary cyst,” “bleb/blebs,” “bulla/bullae,” “atelectasis”
  - Pay particular attention to lung imaging (Chest CT, Chest XR) → use search function for internal radiology notes and for mention of “pneumothorax,” “lung cyst,” “pulmonary cyst,” “bleb/blebs,” “bulla/bullae”
- Manually examine Chest CT radiology reports for mention of “pneumothorax,” “lung cyst,” “pulmonary cyst,” “bleb/blebs,” “bulla/bullae”
- If found:
  - Capture number of pneumothoraces and capture age(s) of diagnosis or, if no age of diagnosis is clear capture the earliest age was documented.
  - For cysts, capture age(s) of diagnosis or, if no age of diagnosis is clear capture the earliest age was documented. If the number of cysts is noted and/or their location, capture that.
- If Pulmonary Cyst code pulled automatically from the chart, look at why that code was identified.

**Renal**

- Search Problem List → any mention of renal cancer
- Use text search function and look for → “renal cancer,” “renal oncocytosis”
- If found → look at surgical pathology and capture pathological diagnoses and age(s) of diagnosis
  - Unilateral, bilateral
  - Multifocal
- Foundation One or Caris Tumor Testing
- If no renal cancer → search for “renal mass”

**Any Cancer History**

- Search problem list and past medical history → any mention of cancer
- Use text search function and look for → “cancer,” “carcinoma”

**Specialists**

- Search for care in certain specialties → Encounters → Filters → Department Specialty
  - Urology
  - Nephrology
  - Dermatology
  - Pulmonology
  - General/Pediatric Genetics
  - Cancer Genetics – Listed under hematology/oncology, so use the search function and search “Genetics”
- Collect if seen

**Family History**

- Search family history tab
- Use search function to search for family history of:
  - Renal cancer
  - Pneumothorax
  - Birt-Hogg-Dubé
  - *FLCN* variant
  - Skin findings– “fibrofolliculoma,” “trichodiscoma”
- Other family history – any other findings of note e.g., renal mass, lung cyst in family member

**Scans**

- Review for any external imaging, outside notes, surgical pathology

**Imaging Review Manual for Participants with *FLCN* Variant**

1. For each patient, the list of imaging studies was reviewed to identify those that might include all or part of the lungs or kidneys, including the following:

- CT of the chest, abdomen, or pelvis
- MRI of the abdomen
- CT and MRI of the thoracic or lumbar spine
- PET-CT (whole body)
- Ultrasound of the kidneys

2. For all studies identified in Step 1, the images were reviewed with attention to the lungs and kidneys.
3. Any relevant findings were noted and then compared to the clinical radiology report within the EHR. Any novel findings were noted as such.
