## Supplemental Table 1 for "Frequency of *FLCN* Loss of Function Variants and Birt-Hogg-Dubé-Associated Phenotypes in a Healthcare System Population"

**Supplemental Table 1: Birt-Hogg-Dubé Syndrome Phenotype-Associated ICD-9/10 Diagnosis Codes**

|  | **ICD-9 Codes** | **ICD-10 Codes** |
| --- | --- | --- |
| **Cutaneous** |  |  |
| Fibrofolliculoma | 216.*, 792.9 | D23.9, R89.6 |
| Trichodiscoma | 215.*, 216.* | D21.9, D23.9 |
| **Pulmonary** |  |  |
| Pulmonary cysts | 518.89, 492.0 | J98.4, J43.9 |
| Spontaneous pneumothorax | 512.0, 512.81, 512.89, 512.8, V12.69 | J93.0, J93.11, J93.83, J93.9 |
| **Renal** |  |  |
| Hybrid oncocytic tumor | - | - |
| Chromophobe renal cell carcinoma | 189.0 | C64.9, C64.1, C64.2 |
| Renal oncocytoma | 223.0 | D30.00. D30.01, D30.02 |
| Renal oncocytosis | - | - |
| Clear cell renal cell carcinoma | 189.0 | C64.9, C64.1, C64.2 |
| Personal h/o malignant neoplasm of kidney | V10.52 | Z85.528 |
| Renal mass | 593.89 | N28.89 |
| **Genetic/Familial Risk** | 759.89, V84.09, V84.89, V19.8 | Q87.89, Z15.09, Z15.89, Z84.89 |
