## Supplemental Table 2 for "Frequency of *FLCN* Loss of Function Variants and Birt-Hogg-Dubé-Associated Phenotypes in a Healthcare System Population"

**Supplemental Table 2. BHD-related phenotypes Captured Using Diagnostic Compared to Manual Review of EHR Data in Variant-Positive Individuals.**

| **Phenotype** | **Manual Chart Review** | **ICD9/10 Diagnosis** |
| --- | --- | --- |
| Spontaneous Pneumothorax | 17.1% (6/35) | 17.1% (6/35) |
| Cystic Lung Disease | 65.7% (23/35) | 20.0% (7/35) |
| Cutaneous Phenotype | 42.9% (15/35) | 8.6% (3/35) |
| Renal Cancer | 2.9% (1/35) | 2.9% (1/35) |
| Renal Mass^a^ | 2.9% (1/34) | 8.8% (3/34) |

^a^Excludes individual with renal cancer diagnosed
