## Supplemental Table 3 for "Frequency of *FLCN* Loss of Function Variants and Birt-Hogg-Dubé-Associated Phenotypes in a Healthcare System Population"

**Supplemental Table 3. Associations Between *FLCN* P/LP Variants and a Diagnosis of BHD-related Phenotypes in the in the First- and Second-Degree Unrelated MyCode Cohort^a^**

| **Phenotype** | **Patient-Participants with P/LP *FLCN* Variant**  **N=28** | **Remaining MyCode Cohort**  **N=** **90,535** | **Odds Ratio (95% CI)** | **p-value^b^** |
| --- | --- | --- | --- | --- |
| **Any Pulmonary Phenotype** | 7 (25%) | 7,103 (7.8%) | 3.93 (1.57-8.83) | **0.03*** |
| Spontaneous Pneumothorax | 6 (21.4%) | 1,723 (1.9%) | 14.76 (5.64-33.48) | **2.66 x 10^-5^*** |
| Pulmonary Cyst | 5 (17.9%) | 5,760 (6.4%) | 3.24 (1.12-7.87) | 0.22 |
| **Cutaneous Phenotype** | 11 (39.3%) | 17,650 (19.5%) | 2.56 (1.17-5.41) | 0.14 |
| **Any Renal Phenotype** | 1 (3.6%) | 2,438 (2.7%) | 1.87 (0.21-7.18) | >0.99 |
| Renal Mass | 0 (0%) | 1,884 (2.1%) | 0.79 (0.01-5.63) | >0.99 |
| Renal Cancer | 1 (3.6%) | 924 (1.0%) | 4.97 (0.55-19.42) | 0.88 |

^a^Ability of ICD-9/10 codes in the EHR to accurately identify BHD-related phenotypes varied between phenotypes.

^b^Bonferroni corrected for multiple testing (7-tests).
